# PheBee: A Graph-Aware System for Scalable, Traceable, and Semantic Phenotyping

**DOI:** 10.64898/2026.05.09.26352812

**Authors:** David M. Gordon, Max Homilius, Austin A. Antoniou, Connor Grannis, Grant E. Lammi, Adam C. Herman, Ashley Kubatko, Bimal P. Chaudhari, Peter White

## Abstract

**Objectives:** Phenotype-driven workflows in clinical and translational research require standardized ontology-based representation, ontology-aware cohort discovery, and provenance inspection for each assertion. Existing approaches optimize either for semantic traversal or scalable batch analytics, but not both. We describe PheBee, a hybrid system that links semantic assertions to scalable evidence storage via a deterministic identifier, preserving provenance while supporting ontology-aware discovery at cohort scale.

**Materials and Methods:** PheBee represents phenotype assertions in a knowledge graph as ontology-linked nodes with clinical modifier context (e.g., negated, family history), and stores supporting evidence records in a scalable row-oriented evidence table for cohort-scale access. The two layers are connected by a deterministic identifier enabling stable joins across repeated ingestions without duplicating high-volume evidence in the graph. We evaluated PheBee using synthetic datasets designed to exercise end-to-end ingestion and query workflows.

**Results:** Functional evaluation validated hierarchical term expansion, qualifier-aware retrieval, duplicate-free assertion handling under re-ingestion, and privacy-conscious management of subjects shared across multiple research projects. At scale (10,000 subjects producing 12M evidence records) PheBee completed ingestion in ∼30 minutes and responded to interactive queries within 6 seconds under concurrent load.

**Discussion:** PheBee exposes a unified API for ontology-aware cohort discovery with hierarchical term expansion, subject-centric retrieval of phenotypes and clinical modifiers, and evidence and provenance queries. Its data model aligns with GA4GH Phenopackets, facilitating interoperability with phenotype exchange standards.

**Conclusion:** By combining ontology-aware semantics with scalable, provenance-bearing evidence storage, PheBee provides a practical open-source foundation for phenotype-driven research workflows that demand both semantic precision and cohort-scale traceability.

**LAY SUMMARY:** Researchers often use “phenotypes” (observable clinical features) to describe individual subjects and find groups of similar subjects. Those phenotypes come from many sources and need both standard terminology and clear evidence for why a phenotype has been associated with a subject.

PheBee is a software system that stores phenotype assertions in a way that supports both “ontology-aware” searching (for example, finding patients with any subtype of a condition) and scalable storage of supporting evidence across large research cohorts. PheBee uses multiple types of data storage so researchers can perform interactive phenotype searches and also store millions of pieces of supporting evidence. A shared identifier connects the two storage layers, so subjects’ phenotypes and their supporting evidence remain linked even as new data is added over time. We evaluated PheBee using fully synthetic (non-patient) data to confirm correct query behavior, evidence traceability, and system performance at large scale.

## BACKGROUND AND SIGNIFICANCE

Phenotype-driven research is governed by a challenging dual-mandate: the simultaneous need for deep semantic awareness and scalable, traceable evidence storage. On one hand, investigators require standardized concepts and ontology-aware querying over resources such as the Human Phenotype Ontology (HPO)[1], to identify cohorts, characterize subjects, and support downstream analysis. On the other hand, because this phenotypic evidence is extracted from heterogeneous sources, including unstructured notes and structured clinical data, investigators also require cohort-scale storage and provenance tracking to audit the origin of each asserted clinical finding. This creates an architectural choice between the semantic intelligence of a knowledge graph and the robust, cohort-scale storage of a data lake, ultimately compromising the efficiency and reproducibility of translational research.

Existing solutions often optimize for only one side of this problem. Phenotype-driven research commonly relies on foundational semantic tools and ontology-backed representations to support consistent terminology, hierarchical reasoning, and reuse across studies. Specifically, HPO and Mondo[2] provide controlled vocabularies for phenotypic abnormalities and diseases, respectively, successfully supporting computational reasoning over complex term relationships. Similarly, data exchange standards like GA4GH Phenopackets provide a standardized schema for exchanging phenotype and disease payloads across tools and institutions[3].

Current large-scale biomedical storage systems integrate phenotype data in various ways. PhenoTips is genomics-focused, providing interfaces for standardized clinical phenotyping and patient-centric curation in diagnostic workflows[4], while systems like i2b2[5] and OHDSI/OMOP[6] allow investigators to build cohorts and impose common data models across massive structured datasets. These lake and warehouse approaches excel at scalable storage and batch analytics, but often lack native ontology traversal and assertion-centric linkage back to clinical context and provenance without supplementary storage or explicit mappings[7,8].

Conversely, pure knowledge graph approaches enable ontology-aware traversal, such as descendant expansion and relationship-centric queries, but can be less efficient for storing and querying high-volume evidence at cohort scale[9–11].

While each of these projects provides significant value to the medical and research communities, there remain opportunities to improve the integration of biomedical knowledge and healthcare data in a way that enhances translational research and clinical care. Together, these gaps between existing tools and clinical research needs motivate a system that supports ontology-aware cohort discovery while retaining traceable evidence records at cohort scale. We developed **PheBee**, a hybrid system that combines a semantic layer for ontology-aware traversal and interactive retrieval with a cohort-scale evidence store for provenance-bearing records.

The key to bridging these two paradigms is a stable, deterministic identifier computed from the subject, ontology term unique identifier, and qualifier context, which acts as an immutable bridge between the graph and the data lake. Building on this unified architecture, PheBee exposes a single API combining ontology-aware cohort discovery with hierarchical term expansion, subject-centric retrieval of phenotypes with their clinical modifiers, and detailed evidence and provenance retrieval. Furthermore, its data model aligns with GA4GH Phenopackets, facilitating interoperability with existing phenotype exchange standards.

## MATERIALS AND METHODS

### Requirements and design goals

We derived requirements from common phenotype-driven workflows in clinical and translational research, including cohort discovery by ontology term, subject-centric review of phenotype assertions, and inspection of supporting evidence and provenance. Although driven by internal pediatric clinical and research workflows, we expect the requirements to generalize to other phenotype-aware domains. We organize them into four themes that drive both the system design and evaluation (**Table 1**):

- Semantic representation and ontology reasoning (R1–R2)
- Interoperability and exchange boundary (R3)
- Provenance, context, and idempotent operations (R4–R6)
- Governance, scalability, and operational robustness (R7–R12)

**Table 1.** PheBee system requirements, organized by design theme. Requirements were derived from phenotype-driven workflows in clinical and translational research and grouped into four themes: semantic representation and ontology reasoning (R1–R2), interoperability and exchange boundary (R3), provenance, context, and idempotent operations (R4–R6), and governance, scalability, and operational robustness (R7–R12). Each requirement informed both system design and the evaluation plan.

| ID | Requirement | Description |
| --- | --- | --- |
| R1 | Standards-based phenotype terms | The system must represent phenotypes/diseases as ontology term IRIs (e.g., HPO, Mondo) and record ontology version metadata so results can be interpreted and reproduced. |
| R2 | Ontology-aware querying | The system must support ontology-aware queries (e.g., descendant/ancestor expansion) for cohort retrieval and summarization. |
| R3 | Interoperable exchange boundary | The system should align with exchange formats such as GA4GH Phenopackets, while separating portable payload exchange from platform concerns like governance, provenance, and operational querying. |
| R4 | Queryable provenance per phenotype assertion | The system must capture and expose provenance for each phenotype/term association, including at minimum an evidence reference, a creator/agent, and timestamps; and (where applicable) text-source pointers (e.g., note ID and span start/end). The system must support filtering/retrieval based on that provenance. |
| R5 | Unified multi-source phenotypes | The system must support both manual curation and automated extraction under the same model, enabling provenance-filterable views (e.g., curated-only vs extracted-only). |
| R6 | Stable, idempotent write operations | The system must provide create/remove operations for core entities (e.g., Subject/Encounter/ClinicalNote/TermLink) with idempotent behavior and predictable identifiers/uniqueness rules. TermLinks must be unique per (*source node, term, context*), where context includes qualifiers such as positive, negated, hypothetical, and family history. |
| R7 | Privacy-conscious multi-project partitioning | The system must maintain a shared subject representation that can be reused across workflows (avoiding duplicate subject records), while keeping project-specific IDs and links isolated per project. This enables complete within-project querying while reducing the risk of unintentionally linking subjects across projects. |
| R8 | Access control and auditability | The system must enforce authorized access to APIs and data, and provide audit-friendly logging for write operations and provenance queries in alignment with institutional governance. |
| R9 | Reliable ingestion and updates | The system must support bulk loads and incremental updates (e.g., ingestion orchestration, load job monitoring, ontology updates, retries) so the stored data can be refreshed safely as upstream data evolve. |
| R10 | Bulk/batch access for analytics and ML | In addition to interactive ontology-aware querying (R2), the system must provide bulk/batch access patterns that support large-scale operations (e.g., cohort-level exports/summaries, feature extraction, and ML training/refresh pipelines). |
| R11 | Scalability and interactive performance | The system must support growth to tens of thousands of patients and millions of rows of phenotypic evidence, including both interactive query workloads and bulk/batch processing. For a defined set of representative interactive queries under expected load, P95 latency should be $\leq 5$ seconds. |
| R12 | Standardized deployment and operations | The system should support repeatable deployment (ideally via infrastructure-as-code) and basic operational support (logging/metrics, integrity checks) appropriate to the current deployment stage. |

### Key terminology

- **Ontology term.** A phenotype or disease concept represented by an Internationalized Resource Identifier (IRI) (e.g., http://purl.obolibrary.org/obo/HP_0002297) (R1). Ontology-aware operations (e.g., descendant expansion) operate over the ontology hierarchy rather than string labels.
- **Assertion.** A claim that an ontology term applies to a subject under a specified qualifier context. In PheBee, assertions are represented as TermLink nodes and are keyed by (subject, term IRI, qualifier context), enabling idempotent ingestion (R6).
- **Qualifiers / contexts.** Contextual modifiers applied to an assertion (e.g., negated, hypothetical, family history). Qualifiers are treated as part of assertion identity for uniqueness and joinability and may be sourced from upstream extractors or curation tools.
- **TermLink.** The PheBee-specific representation of a phenotype assertion in the knowledge graph. Each TermLink node connects a Subject to an ontology term IRI under a specified qualifier context, and carries a deterministic identifier (termlink_id) computed from those three inputs. This identifier associates the TermLink with its supporting records in the evidence layer. TermLinks are unique per (subject, term IRI, qualifier context), enforcing idempotent assertion creation (R6).
- **Idempotent operation.** An operation that produces the same outcome regardless of how many times it is applied. In PheBee, assertion creation is idempotent: submitting the same (subject, term IRI, qualifier context) multiple times results in a single TermLink rather than accumulating duplicates (R6).
- **Evidence record.** A provenance-bearing record supporting an assertion, stored in the evidence layer (R4). Evidence records include termlink_id, subject identifier, optional clinical context identifiers (e.g., encounter/note), creator metadata, timestamps, qualifier type/value pairs, and optional text anchoring (e.g., span offsets) when derived from text.
- **Provenance.** Metadata required to interpret and audit assertions, including creator/agent metadata, timestamps, qualifiers, and links back to supporting evidence and context (R4).
- **Exchange boundary.** The interface at which PheBee maps its internal representation to external exchange formats. At this boundary, PheBee aligns with GA4GH Phenopackets to enable interoperability with other tools and institutions, while keeping internal optimizations for ontology traversal and cohort-scale evidence access separate from the portable payload format (R3).

### Data assumptions and scope

For bulk ingestion of clinical data with full provenance, PheBee assumes upstream systems (extraction pipelines, curation tools, or structured sources) provide phenotype evidence as record-oriented data that can be represented as newline-delimited JSON (JSONL). Records include at minimum an ontology term IRI, a subject identifier, and provenance fields (creator metadata and timestamps); optional fields include qualifiers and text anchoring (e.g., span offsets) when derived from clinical text.

PheBee focuses on representing phenotype assertions and provenance-bearing evidence to support ontology-aware cohort discovery, subject-centric inspection, and cohort-scale analytics. It does not replace upstream extraction or curation systems; instead, it provides repeatable ingestion semantics, stable joins between assertions and evidence, and privacy-conscious multi-project partitioning via shared subject representation with project-scoped identifiers (R7).

### System design

#### Data model and provenance

PheBee represents subjects and phenotype assertions as entities in an RDF (Resource Description Framework) knowledge graph, enabling ontology-aware traversal via linked nodes and edges. High-volume phenotypic evidence and provenance are stored as row-oriented records in a lakehouse-style evidence layer suited to cohort-scale analytics.

Assertions are represented as TermLink nodes that connect a Subject to an ontology term IRI and qualifier context and include termlink_id as a join key to supporting evidence.

Clinical context (e.g., encounter and note identifiers) is retained as optional fields on evidence records, enabling provenance-aware inspection and cohort-scale analytics without requiring those entities to be modeled in the graph. Core entities in the semantic and evidence layers are summarized in **Table 2**.

**Table 2.** Core entities in the PheBee data model. Entities are organized by storage layer. The knowledge graph (upper section) stores shared Subject representations and TermLinks to support ontology-aware traversal and interactive querying. The evidence layer (lower section) stores provenance-bearing evidence records for cohort-scale analytics. Arrows (→) denote primary modeled relationships; queries may traverse edges in either direction, including inverse traversal in SPARQL, the standard query language for RDF graphs.

| Entity Type | Role | Key relationships (high level) |
| --- | --- | --- |
| **Knowledge graph** |  |  |
| **Subject** | Shared subject representation reused across projects | **Subject → ProjectSubjectId**<br>**Subject → TermLink** |
| **Project** | Governance / organizational container used to scope project-specific identifiers |  |
| **ProjectSubjectId** | Binds Subject and Project; stores project-scoped subject identifier | **ProjectSubjectId → Project** |
| **TermLink** | Assertion of evidence connecting a Subject and Ontology term | **TermLink → Ontology term**<br>**TermLink → termlink_id**<br>**TermLink → qualifiers** (e.g. negated, hypothetical, family history) |
| **Evidence layer** |  |  |
| **Evidence** | Evidence/provenance records (e.g., extracted mentions, curated entries), with optional text grounding | **Evidence → termlink_id**<br>**Evidence → clinical context IDs** (subject, encounter, note)<br>**Evidence → creator metadata**<br>**Evidence → qualifiers**<br>**Evidence → text annotation** (span offsets, metadata) |

### Model walkthrough: example phenotype assertions

To illustrate the data model, consider the following synthetic clinical note excerpt:

> *“Family history: Parent diagnosed with dilated cardiomyopathy. Caregiver reports poor exercise tolerance. The patient has a decreased ability to exercise and can’t keep up with peers at recess.”*

In PheBee, phenotype assertions from the note are represented as TermLink nodes attached to the Subject, while encounter/note identifiers and optional text anchoring are recorded on the corresponding evidence rows. Two distinct assertions can be derived:

1. A family history assertion mapped to Dilated cardiomyopathy (HP:0001644), represented as a TermLink with a family history qualifier. One evidence item supports the assertion and includes pointers to the span “dilated cardiomyopathy.”
2. A patient finding mapped to Exercise intolerance (HP:0003546), represented as a TermLink with no qualifiers (positive is the default interpretation). Two evidence items support the assertion and include pointers to the spans “poor exercise tolerance” and “decreased ability to exercise.”

Each evidence row references the associated assertion via termlink_id and carries the provenance and clinical context needed for inspection and downstream analysis.

### Qualifiers and uniqueness

TermLinks may include qualifier type/value pairs that capture assertion context (e.g., negated, hypothetical, family history). While PheBee commonly uses this small set, the model accepts and persists additional qualifier types provided by upstream extraction or curation tools. PheBee treats the qualifier set as part of assertion identity: TermLinks are unique per (subject, term IRI, qualifiers). A deterministic identifier (termlink_id) computed from from these inputs provides a stable join key between the graph assertion and its supporting evidence records:

termlink_id = sha256({subject_iri}|{term_iri}|{comma_joined_sorted_qualifiers})

This strategy hashes false qualifiers equivalently to missing qualifiers, ensuring that new qualifier types can be added to PheBee without invalidating past termlink_id values. If terms are deprecated and replaced, hashes will intentionally continue using the old term_iri to correctly reflect the original term capture, and if semantic links to the replacement term exist, they could be used at query time to consolidate old and new terms into one cohort.

### Intentional scope limitations for initial release

Several aspects of the problem space are intentionally deferred in this initial release, either to downstream applications or to future work:

- Interpretation of conflicting phenotype assertions (e.g., positive and negated mentions for the same subject and term) is left to downstream applications.
- No defined evidence record types beyond text-derived annotations (e.g. structured EHR-derived phenotypes).
- No claims of clinical truth are made as part of this work.

### System architecture

PheBee uses a two-layer architecture that separates semantic assertions from high-volume evidence retention. Phenotype assertions are represented as TermLink nodes in an AWS Neptune-backed knowledge graph to support ontology-aware traversal and interactive queries, while supporting evidence is stored as row-oriented records in an evidence store for cohort-scale analytics and provenance inspection using Apache Iceberg, an open-source table format (**Figure 1**).

**Figure 1.**
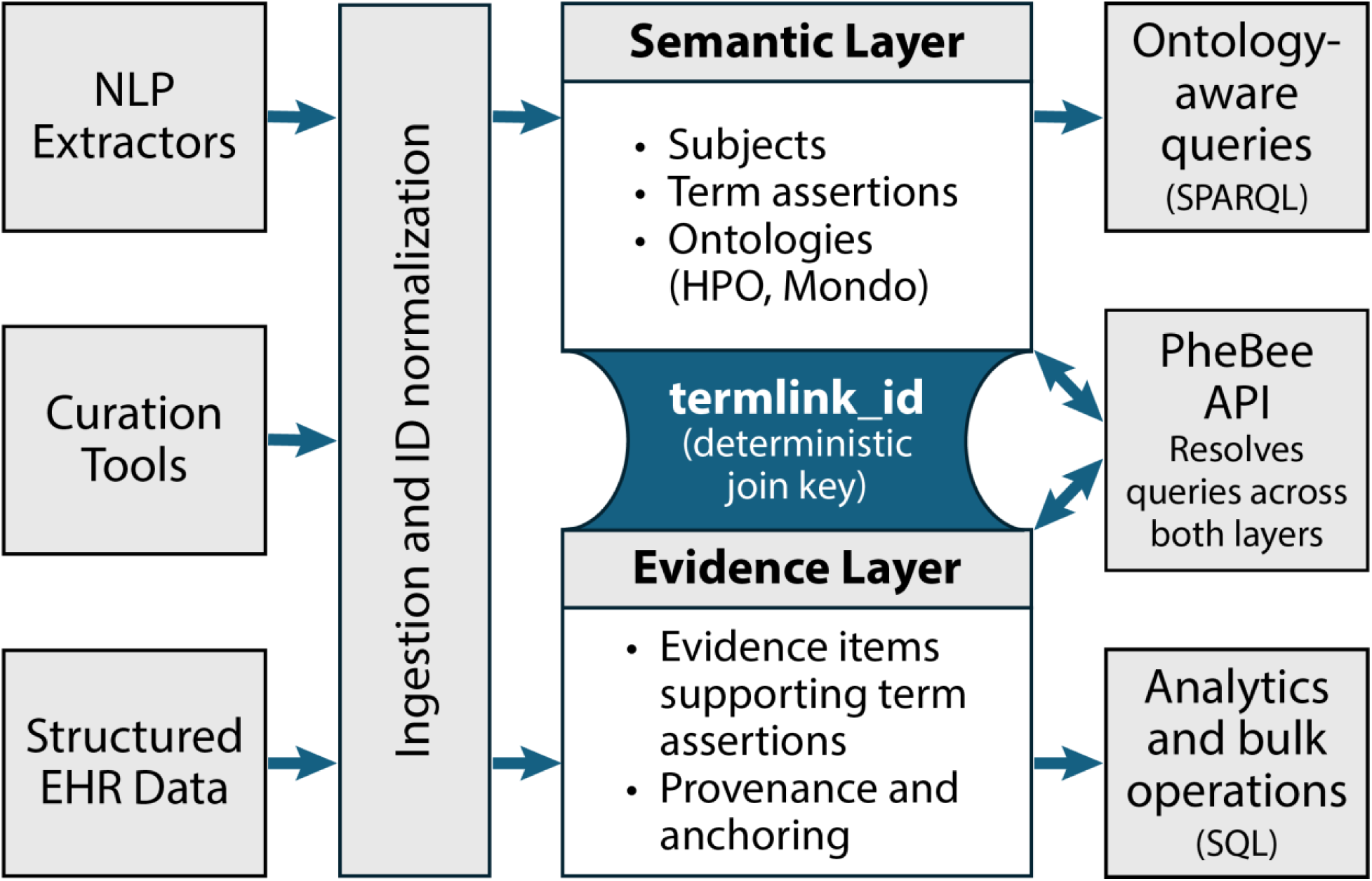
PheBee system architecture. Phenotypic evidence from heterogeneous sources (NLP extractors, curation tools, structured EHR data) is ingested through a normalization pipeline that derives a deterministic join key (termlink_id) for each assertion. Semantic assertions are stored in the knowledge graph layer, supporting ontology-aware traversal and interactive queries via SPARQL. Provenance-bearing evidence records are stored in the evidence layer for cohort-scale analytics and bulk operations via SQL. A unified PheBee API resolves queries across both layers, supporting ontology-aware cohort discovery, subject-centric retrieval, and evidence inspection without requiring callers to interact with each layer directly.

**Figure 1 Alt Text:** Schematic architecture of PheBee showing three phenotypic evidence sources flowing through an ingestion and ID normalization step into two linked storage layers. The semantic layer contains subjects, term assertions, and ontologies such as Human Phenotype Ontology and Mondo. The evidence layer stores evidence items with provenance and anchoring. A deterministic termlink_id connects the layers. The PheBee API resolves queries across both layers, supporting ontology-aware SPARQL queries and SQL-based analytics and bulk operations.

### System components

PheBee comprises a knowledge graph for Subjects and TermLinks, an evidence store for provenance-bearing evidence records, a bulk ingestion workflow that writes evidence and ensures corresponding assertions exist, and an API layer that supports cohort discovery, subject-centric retrieval, and evidence inspection.

### Ingestion and storage

Bulk ingestion reads JSONL evidence records from object storage. For each record, PheBee derives termlink_id from (subject identifier, term IRI, qualifier context), writes the evidence row, and ensures the corresponding TermLink exists. Because TermLinks are keyed by (subject, term IRI, qualifier context), re-ingestion does not create duplicate assertions; additional evidence records accumulate under the same termlink_id.

### Knowledge graph layer

The knowledge graph stores shared Subject representations and the TermLinks needed for interactive query patterns and ontology-aware traversal, while omitting high-volume evidence. Project-scoped identifiers and mappings are isolated using named graphs. During ingestion, derived assertions are serialized using Turtle, an RDF text format, and loaded via the Neptune bulk loader; evidence is resolved through joins on termlink_id.

### Evidence store layer

The evidence store is implemented as an Apache Iceberg table and serves as the primary repository for provenance-bearing evidence. Deployments can optionally target an existing governed Iceberg table. Each row includes termlink_id and record-level provenance and context fields sufficient for audit and downstream analytics, enabling cohort-scale workflows (export, aggregation, feature extraction, ML) without requiring graph-native access for batch computation.

### API and query patterns

PheBee exposes a domain-oriented API for cohort discovery, subject-centric term retrieval (with qualifiers), and evidence/provenance inspection. Clients request subjects, linked terms, or supporting evidence, while the service resolves the appropriate combination of back-end lookups. To support interactive API access patterns, the Iceberg layer also includes materialized views of subject–term associations.

#### Cohort discovery by ontology term

PheBee provides a project-scoped cohort query that lists subjects and optionally filters by an ontology term IRI. When term_iri is provided, include_child_terms (default true) expands to descendants. The query also supports qualifier handling (include_qualified) and cursor-based pagination (limit, cursor) for large result sets, accessed via POST /subjects/query.

If the term_association_source_entity parameter is provided with an identifier representing any entity from the Monarch Knowledge Graph[12], such as a disease, anatomical structure, gene, or other ontology term, PheBee calls the Monarch API to retrieve all associated HPO and Mondo terms. It then searches its own data and returns subjects linked to any of those terms.

#### Subject-centric retrieval of linked terms and evidence

For subject review and downstream feature construction, PheBee provides a subject summary endpoint that returns linked terms, and a term-level drill-down endpoint that returns the underlying TermLinks and associated evidence (resolved via termlink_id). The drill-down accepts optional qualifier filters to distinguish contexts for the same term (e.g., family history vs. patient assertion), accessed via POST /subject (summary) and POST /subject/term-info (term drill-down).

#### Direct API operations

PheBee supports direct operations on core resources to enable curation and evidence management. The API includes endpoints to create and remove evidence and term assertions, and to retrieve evidence by identifier, including creator metadata, timestamps, qualifiers, and optional text-anchoring fields when present. Operational endpoints follow a consistent create/get/remove pattern for core resources.

### Platform context

PheBee is implemented and evaluated on Amazon Web Services (AWS) using managed services, with infrastructure-as-code for repeatable deployment. While AWS-specific in this implementation, the design principles are platform-agnostic.

## Evaluation Methods

### Evaluation setup and synthetic dataset generation

We evaluated PheBee using synthetic datasets generated by a purpose-built automated testing framework, avoiding patient data while exercising end-to-end ingestion and query workflows. The testing framework produces JSONL evidence records matching the bulk import schema and supports configurable cohort “shape” parameters (e.g., number of subjects, terms per subject, evidence records per term, and qualifier distributions). Records include an ontology term IRI (with terminology metadata), subject identifiers, qualifiers, creator metadata, timestamps, and optional text anchoring fields. Datasets were staged in Amazon S3 and ingested through PheBee’s serverless bulk import workflow. Full replication instructions and system configurations are available in **Supplementary Methods (Synthetic Benchmark Dataset Generation).**

### Workloads exercised

Workloads were selected to exercise the requirements in Table 1 (R1–R11). We evaluated bulk ingestion and re-ingestion behavior (R6, R9), interactive cohort discovery and subject-centric retrieval (R2, R4), and cohort-scale batch access via Athena queries over the Iceberg evidence table (R10). We additionally validated Phenopackets-aligned mapping at the exchange boundary (R3) and multi-project partitioning by mapping shared subjects across projects while maintaining project-scoped identifiers (R7).

### Metrics and success criteria

Correctness checks validated cohort retrieval via descendant expansion; expected response structure and evidence count for subject-, term-, and evidence-level retrieval including qualifier context; and idempotent behavior under re-ingestion, consistent with uniqueness per (subject, term IRI, qualifiers) (R1–R7). For performance, we measured end-to-end ingestion time and throughput (records/second) for bulk loads (R9–R11), and response latency for representative interactive endpoints under varying client load (R11), reported as P50 (median) and P95 (95^th^ percentile) latency. Operational practices (R12) are described in the system design and were not directly stress-tested here. All reported metrics use wall-clock elapsed time.

### Experimental configurations

We executed two primary configurations against synthetic data: (i) a functional run at moderate scale to validate correctness across ingestion and query workflows, and (ii) performance runs at larger scale to measure ingestion throughput and P50/P95 interactive latency under varying levels of simultaneous client load. Dataset parameters and results are reported in the Results section; evaluation code and synthetic generation logic are available in the open-source repository.

### AI Disclosure

The authors used generative AI tools (ChatGPT, Anthropic Claude, and AWS Q) to assist with manuscript outlining and revision and with engineering tasks (code review, refactoring, test-suite development). All AI-assisted outputs were reviewed and edited by the authors, who are responsible for the final manuscript and codebase.

## RESULTS

### Functional correctness and requirements validation

Functional evaluation validated key correctness properties across ingestion and query workflows. Descendant expansion supported ontology-aware cohort discovery (R2). Subject-centric retrieval returned linked terms with qualifier context and evidence counts, and term-level drill-down returned expected TermLink identifiers and evidence summaries (R4). Re-ingesting identical evidence did not create duplicate TermLinks, consistent with uniqueness per (subject, term, qualifiers) (R6). Multi-project mapping reused a shared subject representation across projects while keeping project-scoped identifiers isolated in project-scoped queries (R7). Terminology metadata (term_source) was queryable in the Iceberg evidence store when provided (R1), and Phenopacket import/export preserved phenotype and disease identifiers through ingestion and retrieval (R3).

### Bulk ingestion throughput

We evaluated bulk ingestion scalability across five dataset sizes (1,000-100,000 subjects) using synthetic data with configured complexity parameters (150-500 HPO terms per subject, 1-50 evidence items per term link) and 10,000 TermLinks per batch file[13]. These ranges were selected to approximate the upper end of summary statistics observed in an internal Neonatal Intensive Care Unit (NICU) population. Phenotypes were sampled from a fixed HPO version to ensure ontology-aware endpoints exercised real hierarchy traversal. Data were modeled after clinically plausible distributions (qualifier proportions: negated 15%, family 8%, hypothetical 5%, 60% of subjects sampled disease-clustered terms). The 100,000-subject dataset (32.5 million TermLinks, 119.9 million evidence items) completed import in 262.6 minutes at 2,063 TermLinks per second. Import time increased sub-linearly relative to data volume up to 50,000 subjects, then approached linear scaling at 100,000 subjects (**Table 3**).

**Table 3.** Bulk ingestion performance across synthetic dataset sizes. Synthetic datasets were generated with 150–500 HPO terms per subject and 1–50 evidence items per TermLink. Time reflects wall-clock elapsed time for end-to-end ingestion, including writes to both the knowledge graph and evidence layers. Throughput is reported separately for TermLinks and evidence items per second.

| Subjects | TermLinks | Evidence | Time (min) | TermLinks/sec | Evidence/sec |
| --- | --- | --- | --- | --- | --- |
| 1000 | 329,240 | 1,214,327 | 12.8 | 428 | 1,577 |
| 5000 | 1,624,945 | 5,994,039 | 22.6 | 1,197 | 4,416 |
| 10000 | 3,251,666 | 11,993,639 | 34.9 | 1,552 | 5,723 |
| 50000 | 16,232,032 | 59,878,126 | 118.4 | 2,285 | 8,428 |
| 100000 | 32,516,163 | 119,945,659 | 262.6 | 2,063 | 7,613 |

### Interactive API performance at scale

We evaluated interactive API latency across each dataset size and varied client-side concurrency levels of 1, 10, and 25 simultaneous clients (c=1, c=10, c=25). For each configuration, we issued 100 requests per workflow across a set of common workflow operations (**Supplementary Methods: Interactive Performance Workload Definitions**), repeated the procedure three times, and report the median of replicate medians for P50 and P95 to reduce run-to-run variability due to transient environmental factors. Workflow times reflect call-to-response latency, not solely database performance.

At 50,000 subjects (Figure 2A), most workflows exhibited P95 latencies of approximately 1 to 6 seconds at concurrency 1, with the paginated large cohort workflow showing the highest latency. Increasing concurrency had relatively little effect on P50 latency for most workflows; workflows involving unfiltered cohort retrieval showed the largest P95 increases at concurrency 25. Similar patterns were observed at dataset sizes of 1,000, 5,000, 10,000, and 100,000 subjects (**Supplementary Figure S1**).

**Figure 2.**
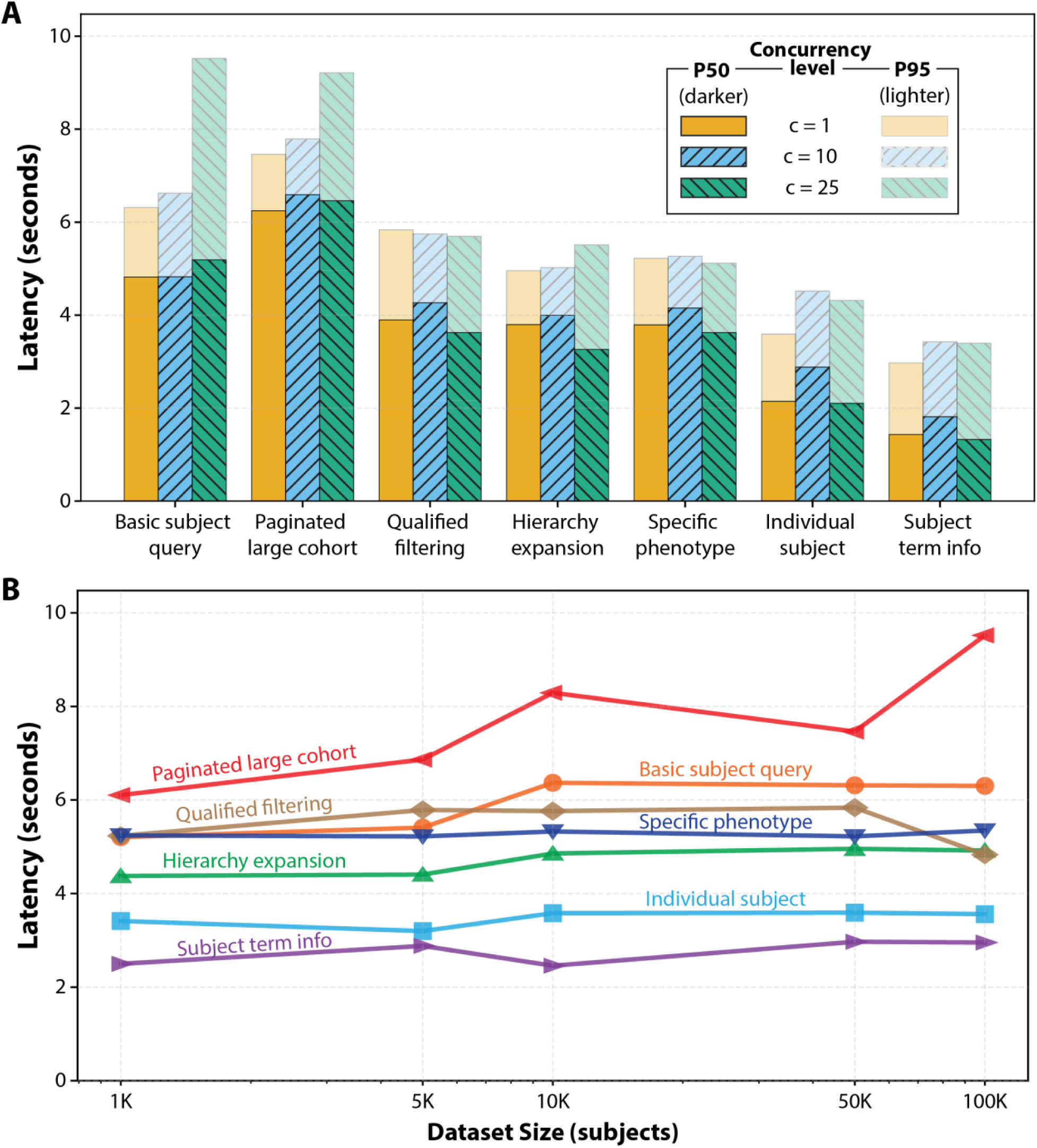
**Interactive API latency scaling under varying concurrency and dataset size**. (A) P50 (median, darker bars) and P95 (95th percentile, lighter bars) latency for each workflow at 50,000 subjects, at concurrency levels of 1, 10, and 25 simultaneous clients. (B) P95 latency across dataset sizes of 1K–100K subjects at concurrency 1. For both panels, 100 requests per workflow were issued per configuration; values are the median of three replicate medians to reduce run-to-run variability.

When concurrency was held constant at one client and dataset size was increased (Figure 2B), P95 latency remained relatively stable for most workflows; unfiltered cohort retrieval workflows showed the largest increases. This pattern was consistent across P50 and P95 measurements as concurrency increased from 1 to 10 and 25 simultaneous clients. (**Supplementary Figure S2**).

**Figure 2 Alt Text:** Two-panel performance figure summarizing PheBee API latency. Panel A shows median and 95th percentile latencies for seven query workflows at concurrency levels of 1, 10, and 25 clients on a 50,000-subject dataset. Most subject-specific and phenotype-filtered workflows show modest latency changes with higher concurrency, while basic subject query and paginated large cohort workflows have the highest 95th percentile latencies, remaining under 10 seconds.

Panel B shows 95th percentile latency across 1,000 to 100,000 subjects at concurrency 1, with paginated large cohort increasing most as dataset size grows, while other workflows remain comparatively stable.

### Batch analytics validation via Athena

Athena queries over the Iceberg evidence table confirmed that the evidence store supports cohort-scale summarization workflows, including term frequency counts and per-subject feature aggregation (R10). Queries also validated that expected context, provenance, and ontology metadata fields were correctly retained and queryable (R1). Together these results confirm that batch analytics workflows can operate directly over the evidence layer without requiring graph-native access.

## DISCUSSION

### Interpretation

PheBee was designed to support phenotype-driven research workflows that require standardized, ontology-linked representation and traceable evidence. The results indicate that a two-layer approach linked by a stable identifier can support semantically aware cohort discovery, subject-centric review, and provenance inspection without requiring all workloads to operate directly against a knowledge graph.

### Tradeoffs of the hybrid design

The hybrid design separates ontology-aware traversal and semantic querying (knowledge graph layer) from cohort-scale evidence retention and batch computation (evidence store). This avoids storing high-volume evidence in the graph and enables analytics workflows to operate directly over the evidence store when appropriate. Tradeoffs include operational complexity of maintaining two persistence layers, the need for a stable linkage mechanism (termlink_id), and multi-step retrieval for query patterns that span ontology-aware traversal and evidence filtering. The boundary between “assertion” and “evidence detail” may evolve as additional evidence types and use cases are integrated.

### Limitations

Evaluation was performed on synthetic datasets generated by a test framework rather than real patient data. While appropriate for validating system behavior without privacy risk, synthetic data may not capture real-world distributions (e.g., term frequency skew, encounter/note patterns, and heterogeneity of upstream extraction). Performance measurements are also sensitive to dataset shape and workload mix. The current implementation requires AWS managed services; support for other environments would require additional development. Finally, the evaluation emphasized representative correctness checks and baseline performance rather than exhaustive workload coverage or long-running stability testing.

### Performance considerations

In the scale configuration, P95 interactive latency exceeded the ≤5-second target under higher concurrency, particularly for unfiltered cohort retrieval. We treat these measurements as a performance baseline for the current implementation and configuration rather than an inherent limitation of the hybrid approach; both layers offer targeted optimization opportunities. In the semantic layer, candidates include SPARQL query tuning and Neptune instance sizing and caching or alternate graph database products. In the API layer, response caching, tighter pagination, and summary-first responses with on-demand drill-down are likely to reduce latency. Performance on real-world clinical data will be evaluated to inform further optimization.

### Interoperability and external data integration

PheBee is designed to interoperate with external resources at multiple levels: ontology currency, knowledge graph integration, and data exchange standards.

HPO and Mondo ontologies are installed and updated automatically when new releases occur, keeping phenotype and disease concepts synchronized with current curation standards. Building on this, PheBee accepts any identifier recognized by the Monarch Knowledge Graph, which integrates data from more than 35 sources spanning gene expression, variation, and disease associations. By integrating with the Monarch API, PheBee can expand a simple concept (for example, “Second Heart Field”) into a set of HPO and Mondo terms derived from those curated associations, then search its own dataset for matching subjects, offering semantically rich cohort identification.

A growing number of systems export patient profiles as GA4GH Phenopackets. PheBee supports this standard for both import and export: records from clinical data entry tools or curated collections such as the Phenopacket Store[14] can be ingested directly, while cohorts assembled in PheBee can be exported as Phenopackets for downstream services.

Together, these integrations position PheBee as a bridge between internal clinical phenotyping and the broader ecosystem of community-curated biomedical knowledge.

### Real-world validation

PheBee is currently deployed as part of an institutional pilot project, ingesting NLP-extracted phenotypic assertions from clinical notes in the level IV NICU. This highest-acuity NICU level serves the most critically ill neonates, and the resulting phenotype data support ongoing research.

Preliminary metrics from this deployment demonstrate that the system can operate at production scale (45K total level III+IV NICU subjects, 9M TermLinks, 90M evidence items, ∼5K-10K evidence items imported nightly). The current pilot uses the Subject Term Info workflow to characterize patients; observed latency is modestly higher than the synthetic 50K benchmark under similar load (concurrency ∼10; pilot p50=2.4s, p95=4.1s vs benchmark p50=1.8s, p95=3.4s). A full analysis of that implementation will be reported separately.

### Governance and operational considerations

PheBee targets environments where governance and project boundaries matter. Its model reuses a shared subject representation while scoping project-specific identifiers and mappings to reduce cross-project linkage risk. Fine-grained authorization policies are deployment-specific and were not evaluated here, but the partitioned design supports enforcement at the API layer. Provenance-bearing evidence records (creator metadata, timestamps, qualifiers, optional text anchoring) support review and auditing, and idempotent assertion semantics reduce duplicates and preserve stable joins over time. The AWS implementation supports common operational needs such as authorized API access, centralized logging, and repeatable deployment.

## CONCLUSION

PheBee supports phenotype-aware workflows that require term representation and auditable evidence retention. It uses a hybrid design: assertions are stored in a knowledge graph layer for ontology-aware traversal and interactive queries, while provenance-bearing evidence is retained in a batch-oriented store. A deterministic identifier (termlink_id) links the layers, enabling stable joins across repeated ingestions without duplicating evidence in the graph.

Using synthetic datasets, we validated ontology-based cohort retrieval via descendant expansion, qualifier-aware subject characterization, provenance-preserving evidence retrieval, and interoperability. Scale runs further characterized end-to-end ingestion and interactive query behavior at cohort volumes typical of phenotype-driven pipelines.

Future work falls into two broad areas. In the near term, we will standardize the evidence payload profile (including qualifiers, creator metadata, and text anchoring conventions) and characterize real-world scale and usage patterns to guide performance optimization. Longer term, we will expand beyond text-derived annotations to additional evidence modalities (e.g., structured laboratory sources and LOINC-derived phenotypes) and enrich semantic context by integrating additional ontologies and exposing further graph operations.

By combining ontology-aware semantics with scalable, provenance-bearing evidence storage, PheBee provides a practical open-source foundation for phenotype-driven research workflows that demand both semantic precision and cohort-scale traceability.

## DECLARATIONS

### Ethics approval and consent to participate

This project did not involve any human subjects or real clinical records; all evaluation used fully synthetic datasets generated algorithmically from summary statistics and publicly available ontologies. No protected health information was accessed at any stage, and Institutional Review Board approval was not required.

### Consent for publication

Not applicable.

### Availability of data and materials

Synthetic dataset generation logic and evaluation workloads are included in the project repository. Synthetic datasets used for evaluation are available in Zenodo (DOI: https://doi.org/10.5281/zenodo.19698733)[13].

### Availability of code

PheBee is available as open-source software through GitHub (https://www.github.com/nch-cloud/phebee) and archived in Zenodo (https://doi.org/10.5281/zenodo.19769737)[15]. The repository includes infrastructure-as-code templates, API definitions, and the evaluation harness used in this manuscript.

### Authors’ contributions

David M. Gordon (Conceptualization, Methodology, Software, Validation, Investigation, Formal analysis, Data curation, Writing—original draft, Writing—review & editing), Max Homilius (Methodology, Software, Investigation), Austin A. Antoniou (Methodology, Investigation), Connor Grannis (Methodology, Software, Investigation), Grant E. Lammi (Software, Resources), Adam C. Herman (Software, Resources), Ashley Kubatko (Project administration, Supervision, Resources), Bimal P. Chaudhari (Conceptualization, Resources, Supervision, Funding acquisition, Writing—review & editing), and Peter White (Conceptualization, Supervision, Funding acquisition, Project administration, Writing—original draft, Writing—review & editing). All authors reviewed and approved the final manuscript.

## Data Availability

Synthetic dataset generation logic and evaluation workloads are included in the project repository. Synthetic datasets used for evaluation are available in Zenodo (DOI: https://doi.org/10.5281/zenodo.19698733). PheBee is available as open-source software through GitHub (https://www.github.com/nch-cloud/phebee) and archived in Zenodo (https://doi.org/10.5281/zenodo.19769737). The repository includes infrastructure-as-code templates, API definitions, and the evaluation harness used in this manuscript.

https://doi.org/10.5281/zenodo.19698733

https://doi.org/10.5281/zenodo.19769737

## Acknowledgements

We thank the Nationwide Children’s Hospital Foundation Pediatric Innovation Fund for generously supporting this project. We thank our colleagues in the Office of Data Sciences at Nationwide Children’s Hospital for their technical support, scientific discussions, and feedback that informed the development of PheBee. We also thank the Institute for Genomic Medicine at Nationwide Children’s Hospital, which has supported our phenotyping work through scientific and requirements feedback. Finally, we acknowledge the GA4GH, Monarch Initiative, Robinson lab, and Haendel lab for their foundational contributions to phenotype and disease standardization and interoperability.

## REFERENCES

1 Robinson PN, Köhler S, Bauer S, et al. The Human Phenotype Ontology: a tool for annotating and analyzing human hereditary disease. Am J Hum Genet. 2008;83:610–5. doi: 10.1016/j.ajhg.2008.09.017

2 Vasilevsky NA, Toro S, Matentzoglu N, et al. Mondo: Integrating Disease Terminology Across Communities. Genetics. 2025;iyaf215. doi: 10.1093/genetics/iyaf215

3 Jacobsen JOB, Baudis M, Baynam GS, et al. The GA4GH Phenopacket schema defines a computable representation of clinical data. Nat Biotechnol. 2022;40:817–20. doi: 10.1038/s41587-022-01357-4

4 Girdea M, Dumitriu S, Fiume M, et al. PhenoTips: patient phenotyping software for clinical and research use. Hum Mutat. 2013;34:1057–65. doi: 10.1002/humu.22347

5 Murphy SN, Weber G, Mendis M, et al. Serving the enterprise and beyond with informatics for integrating biology and the bedside (i2b2). J Am Med Inform Assoc. 2010;17:124–30. doi: 10.1136/jamia.2009.000893

6 Hripcsak G, Duke JD, Shah NH, et al. Observational Health Data Sciences and Informatics (OHDSI): Opportunities for Observational Researchers. Stud Health Technol Inform. 2015;216:574–8.

7 Callahan TJ, Stefanski AL, Wyrwa JM, et al. Ontologizing health systems data at scale: making translational discovery a reality. NPJ Digit Med. 2023;6:89. doi: 10.1038/s41746-023-00830-x

8 Post A, Chappidi N, Gunda D, et al. A Method for EHR Phenotype Management in an i2b2 Data Warehouse. AMIA Jt Summits Transl Sci Proc. 2019;2019:92–101.

9 Hölsch J, Schmidt T, Grossniklaus M. On the Performance of Analytical and Pattern Matching Graph Queries in Neo4j and a Relational Database. Proceedings of the GraphQ Workshop, EDBT/ICDT 2017 Joint Conference. Venice, Italy 2017.

10 Cheng Y, Ding P, Wang T, et al. Which Category Is Better: Benchmarking Relational and Graph Database Management Systems. Data Sci Eng. 2019;4:309–22. doi: 10.1007/s41019-019-00110-3

11 Timón-Reina S, Rincón M, Martínez-Tomás R. An overview of graph databases and their applications in the biomedical domain. Database (Oxford). 2021;2021:baab026. doi: 10.1093/database/baab026

12 Putman TE, Schaper K, Matentzoglu N, et al. The Monarch Initiative in 2024: an analytic platform integrating phenotypes, genes and diseases across species. Nucleic Acids Res. 2024;52:D938–49. doi: 10.1093/nar/gkad1082

13 Gordon D. PheBee Performance Benchmarks: Synthetic Subject-Phenotype Datasets. Zenodo 2026. doi: 10.5281/zenodo.19698733

14 Danis D, Bamshad MJ, Bridges Y, et al. A corpus of GA4GH Phenopackets: case-level phenotyping for genomic diagnostics and discovery. medRxiv. 2024;2024.05.29.24308104. doi: 10.1101/2024.05.29.24308104

15 Gordon D, Homilius M. PheBee. Zenodo 2026. doi: 10.5281/zenodo.19769737

