## Supplementary Methods for "PheBee: A Graph-Aware System for Scalable, Traceable, and Semantic Phenotyping"

### SYNTHETIC BENCHMARK DATASET GENERATION

### Purpose and Scope

Synthetic datasets were generated to enable reproducible performance benchmarking while protecting patient privacy. These benchmarks address three key research questions: (1) bulk ingestion throughput across varying cohort sizes, (2) API query latency under concurrent load, and (3) scalability characteristics of Iceberg analytical table partitioning strategies. Synthetic data enable rigorous testing at scales exceeding available de-identified cohorts while maintaining statistical fidelity to real-world phenotyping patterns.

### Real-World Data Reference

Synthetic dataset parameters were calibrated against summary statistics from an internal NICU phenotyping cohort (N=45,228 subjects, 90.4M evidence records) collected through automated NLP extraction and manual curation. Key distributional characteristics matched in the synthetic data include:

- **Terms per subject**: Production median=114, p75=183, p90=281 (synthetic target: 150-500, representing deeply phenotyped rare disease patients)
- **Evidence items per term link**: Production median=2, p75=6, p90=18, p95=37 (synthetic: 1-50)
- **Qualifier prevalence**: Negated findings (15%), family history (8%), hypothetical diagnoses (5%)
- **Term frequency distributions**: Common phenotypes (70% of assignments) vs. rare findings (30%), derived from production term frequency analysis

### Synthetic Data Generation Process

Benchmark datasets are generated via `generate_benchmark_dataset.py` `(tests/integration/performance/)` using the following algorithm:

**Step 1: Term Universe Construction**

- Parse HPO ontology (`hp.obo`) into searchable JSON index via `generate_hpo_terms_json.py`
- Classify terms as "common" (top 30% by frequency) or "rare" based on optional prevalence CSV. For manuscript benchmarks, prevalence was derived from real world summary statistics on the same NICU cohort
- Extract hierarchical relationships for descendant expansion queries

**Step 2: Subject Generation**

- Generate N unique subject IDs (UUID4 format)
- Assign 60% of subjects to disease clusters (cardiomyopathy, epilepsy, metabolic, oncology, rare dysmorphic) with co-occurring anchor terms
- Remaining 40% receive terms via simple random sampling

**Step 3: Term Assignment**

- For each subject, sample M terms (uniform distribution between min_terms and max_terms)
- For clustered subjects: ensure anchor term inclusion (60% probability), then sample related terms from cluster-specific pool
- Weight sampling by frequency: 70% common terms, 30% rare terms (when prevalence data available)

**Step 4: Qualifier Assignment**

- Assign context qualifiers probabilistically: negated (15%), family_history (8%), hypothetical (5%), unqualified (72%)

**Step 5: Evidence Generation**

- For each term link, sample K evidence items (uniform distribution between min_evidence and max_evidence)
- Weight evidence count by clinical importance: chief complaints (5-12), active problems (2-6), history (1-3), incidental (1-2)
- Generate provenance metadata: creator ID/type, note timestamp (2023-2024 range), specialty attribution

**Step 6: Batch File Creation**

- Split records into 10,000-record NDJSON batch files
- Add row_num (global position) and batch_id fields for traceability

All generation for this manuscript uses fixed random seed (default: 42) for reproducibility. Implementation details available in `tests/integration/performance/conftest.py` (functions: `generate_scale_dataset`, `generate_disease_clusters`).

### Synthetic Dataset Characteristics

Benchmark datasets at five scales (1K, 10K, 100K subjects) exhibit the following summary statistics (10K dataset shown):

**Dataset Composition:**

- Subjects: 10,000
- Term links (records): ~2.5M (mean: 250 terms/subject)
- Total evidence items: ~30M (mean: 12 evidence/term link)
- Unique HPO terms: ~1,200 (1.5% of HPO vocabulary)

**Distributional Fidelity** (Compared to Production p75-p95 Range):

- Terms per subject: min=150, max=500 (production: p75=183, p90=281) ✓ Conservative upper bound
- Evidence per term link: min=1, max=50 (production: p75=6, p95=37) ✓ Captures heavy documentation tail
- Term frequency profile: Pearson r=0.89 with production frequency distribution

**Qualifier Distribution** (Target vs. Observed):

- Negated: 15% target → 14.8% observed
- Family history: 8% target → 8.1% observed
- Hypothetical: 5% target → 5.0% observed

### Performance Evaluation Setup

**Infrastructure:**

- AWS Region: us-east-2
- Compute: AWS Lambda (configuration varies by task, available in `template.yaml`), EMR Serverless (emr-7.10.0, default application settings)
- Storage: Apache Iceberg on S3 (Parquet format), Neptune graph database (1 node, db.r5.large)
- Standard versions:
  - HPO: v2026-01-08
  - Phenopackets: v2

**Bulk Import Protocol:**

- Batch size: 10,000 records per NDJSON file
- Concurrency: 25 parallel Step Functions Map workers
- Timeout: 6 hours

### Data Formats And Accessibility

NDJSON Record Schema:


```
{
  "project_id": "phebee-benchmark-10000subj-seed42",
  "project_subject_id": "subject-0001",
  "term_iri": "http://purl.obolibrary.org/obo/HP_0001250",
  "qualifiers": ["negated"],
  "evidence": [
    {
      "evidence_id": "...",
      "created_timestamp": "2024-05-15T14:32:00Z",
      "evidence_creator_id": "nlp-pipeline-v2.1",
      "evidence_creator_type": "automated",
      "note_type": "progress_note",
      "provider_type": "physician",
      "author_specialty": "neurology"
    }
  ],
  "term_source": {
    "source": "hpo",
    "version": "2024-04-26",
    "iri": "http://purl.obolibrary.org/obo/hp.owl"
  },
  "row_num": 12345,
  "batch_id": 1
}
```


**Reproduction Instructions:**

*Download HPO ontology*

```
curl -L http://purl.obolibrary.org/obo/hp.obo -o data/hp.obo
```

*Generate term index*

```
python generate_hpo_terms_json.py --obo data/hp.obo --out
data/hpo_terms.json
```

*Set parameters*

```
export PHEBEE_EVAL_TERMS_JSON_PATH="data/hpo_terms.json"
export PHEBEE_EVAL_SEED=42
export PHEBEE_EVAL_SCALE_SUBJECTS=10000
```

*Generate benchmark dataset*

```
python generate_benchmark_dataset.py
```

*Output:*

```
data/benchmark/10000-subjects-seed42/
```

**Benchmark datasets available at:** [https://doi.org/10.5281/zenodo.19698733] (includes `metadata.json`, `README.md`, and `batches/` directory).

### Limitations and Realism

**Captured Characteristics:**

- Term frequency distributions matching production
- Disease clustering patterns reflecting real comorbidity networks
- Longitudinal evidence accumulation (temporally distributed annotations)
- Specialty-appropriate attribution (e.g., cardiac phenotypes → cardiology)

**Uncaptured Characteristics:**

- Missing data: Synthetic records have complete metadata
- Textual noise: Evidence text is template-generated
- Temporal patterns: Synthetic timestamps are uniformly distributed; real clinical documentation shows episodic clustering around acute events
- Co-occurrence networks: Disease clustering uses 5 hand-curated phenotype sets; production data exhibits richer comorbidity patterns learned from 1,200+ diagnosis codes
- Long-tail behavior: Synthetic max (500 terms/subject) represents complex patients, but is conservative vs. production max (1,113 terms); extremely complex patients underrepresented

**Impact on Performance Metrics:**

- Import throughput: Synthetic data likely realistic (structured data, varied dataset benchmark scales up to 100K subjects, real clinical data likely to be smaller incremental loads)
- Query latency: Representative for median-complexity patients; may underestimate p99 tail latency for extreme outliers
- Materialization performance: Realistic for target use case (deeply phenotyped research cohorts)

#### Reproducibility

All performance benchmarks can be reproduced using the provided codebase and datasets:

**Quick Reproduction (using pre-generated data):**

1. Download benchmark dataset from Zenodo
[https://doi.org/10.5281/zenodo.19698733]

2. Set environment

```
export PHEBEE_EVAL_SCALE=1
export PHEBEE_EVAL_TERMS_JSON_PATH="data/hpo_terms.json"
export PHEBEE_EVAL_BENCHMARK_DIR="data/benchmark/10000-subjects-seed42"
```

3. Deploy PheBee stack

```
sam build && sam deploy --config-env integration-test
```

4. Run performance tests

```
pytest -v
tests/integration/performance/test_import_performance.py
pytest -v
tests/integration/performance/test_evaluation_perf_scale.py
```

**Full Reproduction (regenerating data):**

- Follow instructions in Section 6 to generate fresh dataset with seed=42
- Ensure HPO version matches (2024-04-26, or specify in `PHEBEE_EVAL_TERMS_JSON_PATH`)
- All tests use fixed random seed (configured via `PHEBEE_EVAL_SEED`) for deterministic sampling

**Configuration Files:**

- `tests/integration/performance/README.md`: Complete setup documentation
- `.phebee-test-stack`: Stack name persistence for test suite

**Hardware Requirements:**

- AWS account with sufficient quotas (50 concurrent Lambda invocations, 200GB S3 storage)
- Local machine: 8GB RAM, 20GB disk for dataset generation

**Estimated Runtime:**

- Dataset generation (10K subjects): ~5 minutes
- Stack deployment: ~20 minutes
- Import performance test (10K subjects): ~35 minutes
- API latency test (700 queries @ 25 workers): ~8 minutes

### INTERACTIVE PERFORMANCE WORKLOAD DEFINITIONS

#### 1) Basic Subjects Query

- **Pattern:** Simple project query with a small limit
- **Use case:** Browsing subjects in a project
- **API call:** POST /subjects/query
- **Parameters:** project_id, limit=10
- **Exercises:** Baseline routing and project-scoped filtering

#### 2) Individual Subject

- **Pattern:** Single subject detail lookup
- **Use case:** Patient phenotype characterization
- **API call:** POST /subject
- **Parameters:** project_subject_iri (rotates through available subjects)
- **Exercises:** Point lookup path for subject-centric retrieval

#### 3) Hierarchy Expansion

- **Pattern:** Descendant expansion with ontology traversal
- **Use case:** "All subjects with cardiovascular conditions" or similar broad query
- **API call:** POST /subjects/query
- **Parameters:** term_iri=HP:0001626 (Abnormality of cardiovascular system), include_child_terms=true
- **Exercises:** Ontology traversal and expanded cohort resolution, caching of term descendant retrieval on repeated query of same term

#### 4) Qualified Filtering

- **Pattern:** Exclude qualified contexts (e.g., negated/family history/hypothetical)
- **Use case:** Clinical queries requiring "confirmed findings only" views
- **API call:** POST /subjects/query
- **Parameters:** term_iri, include_qualified=false
- **Exercises:** Qualifier-aware filtering behavior at scale

**5) Specific Phenotype**

- **Pattern:** Direct term query without hierarchy expansion
- **Use case:** Exact term matching for research queries
- **API call:** POST /subjects/query
- **Parameters:** term_iri (random sample of existing terms, no descendant expansion; include_child_terms=false)
- **Exercises:** Exact-match term-level retrieval without ontology traversal

**6) Paginated Large Cohort**

- **Pattern:** Large result set with cursor-based pagination
- **Use case:** Broad cohort queries returning many subjects
- **API call(s):** POST /subjects/query (repeated if next_cursor is returned)
- **Parameters:** limit=50, follow next_cursor when present
- **Exercises:** Pagination logic, large result handling, repeated-call behavior

**7) Subject Term Info**

- **Pattern:** Detailed subject–term evidence retrieval
- **Use case:** Curator workflows inspecting supporting evidence for a phenotype
- **API call:** POST /subject/term-info
- **Parameters:** subject_id, term_iri (and optional qualifier filter, if present)
- **Exercises:** Drill-down retrieval and evidence resolution for a specific assertion
