## Supplementary Figure S1 for "PheBee: A Graph-Aware System for Scalable, Traceable, and Semantic Phenotyping"

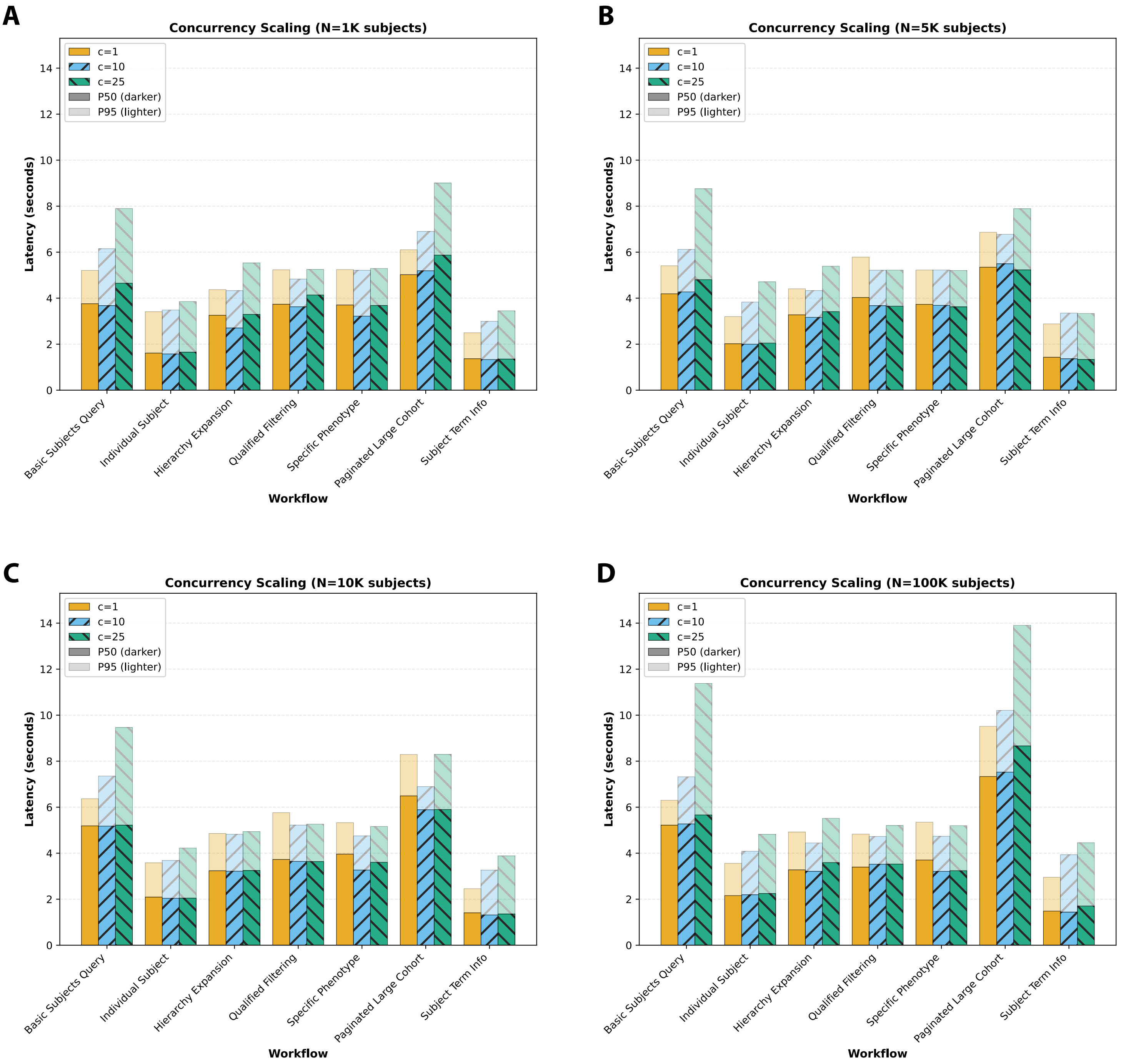


**Supplementary Figure S1.** PheBee API latency by workflow and concurrency across dataset sizes. P50 (median, darker bars) and P95 (95th percentile, lighter bars) latency are shown for seven interactive query workflows at concurrency levels of 1, 10, and 25 simultaneous clients. Panels show performance at dataset sizes of 1,000 subjects (A), 5,000 subjects (B), 10,000 subjects (C), and 100,000 subjects (D). Across dataset sizes, most subject-specific and phenotype-filtered workflows showed modest changes in latency as concurrency increased, whereas broad cohort retrieval workflows, particularly basic subject query and paginated large cohort, showed the largest increases in P95 latency. For each configuration, 100 requests were issued per workflow; plotted values represent the median of three replicate medians.
