## Supplementary Figure S2 for "PheBee: A Graph-Aware System for Scalable, Traceable, and Semantic Phenotyping"

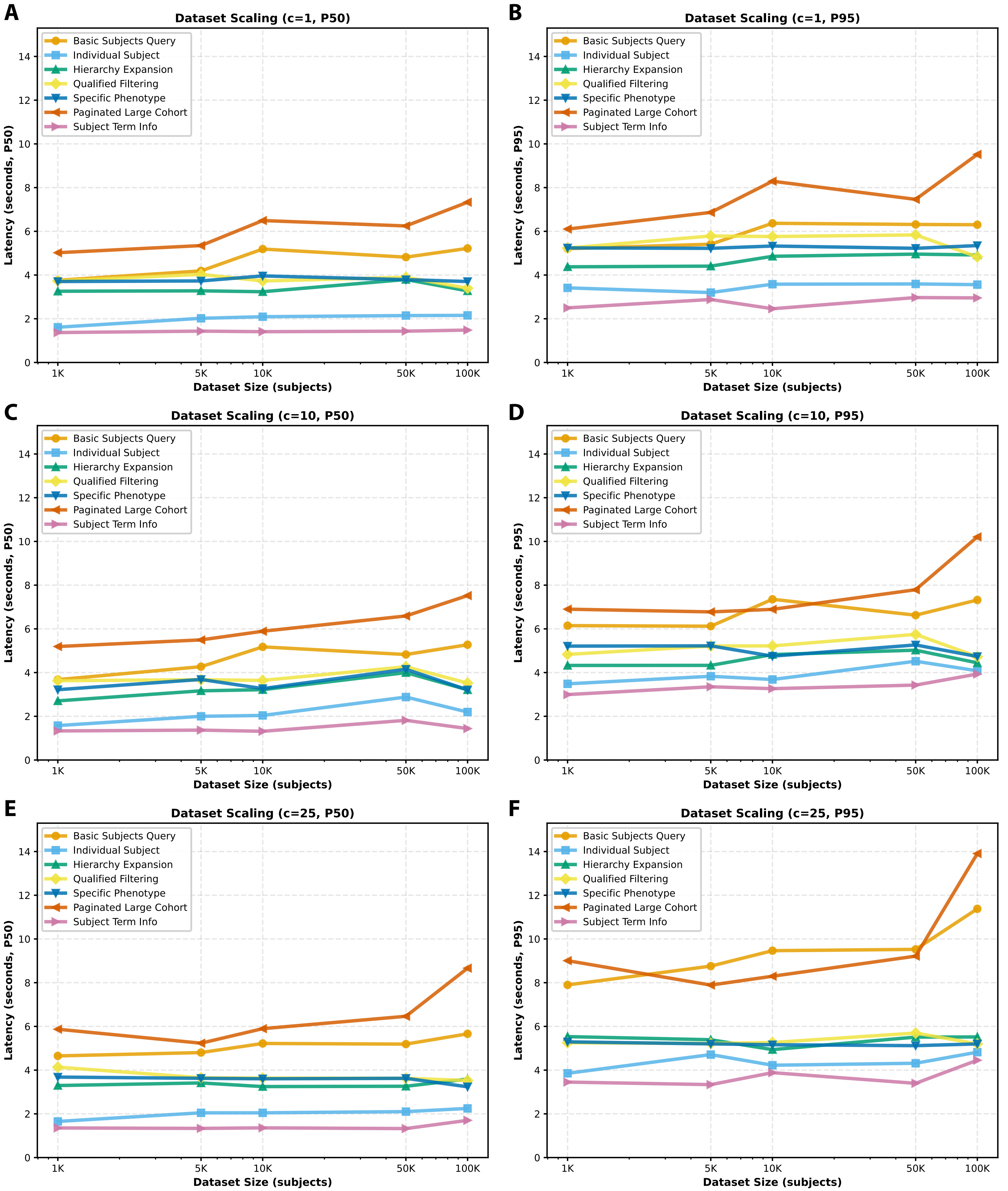


**Supplementary Figure S2.** PheBee API latency scaling by dataset size, concurrency, and latency percentile. Latency was measured across seven interactive query workflows as dataset size increased from 1,000 to 100,000 subjects. Panels show median latency (P50) and 95th percentile latency (P95) at concurrency levels of one client (A, B), 10 simultaneous clients (C, D), and 25 simultaneous clients (E, F). Panel B is identical to Figure 2B. Across concurrency settings, most subject-specific and phenotype-filtered workflows showed relatively stable latency as dataset size increased, whereas broad cohort retrieval workflows, particularly paginated large cohort queries, showed the largest increases. For each configuration, 100 requests were issued per workflow; plotted values represent the median of three replicate medians.
